# Metabolic Disturbances, Hemoglobin A1c, and Social Cognition Impairment in Schizophrenia Spectrum Disorders

**DOI:** 10.1101/2022.04.01.22273310

**Authors:** Sunny X. Tang, Lindsay D. Oliver, Katrin Hänsel, Pamela DeRosse, Majnu John, Ammar Khairullah, James M. Gold, Robert W. Buchanan, Aristotle Voineskos, Anil K. Malhotra

## Abstract

Social cognitive impairments are core features of schizophrenia spectrum disorders (SSD) and are associated with greater functional impairment and decreased quality of life. Metabolic disturbances have been related to greater impairment in general neurocognition, but their relationship to social cognition has not been previously reported. In this study, metabolic measures and social cognition were assessed in 245 participants with SSD and 165 healthy comparison subjects (HC), excluding those with hemoglobin A1c (HA1c)>6.5%. Tasks assessed emotion processing, theory of mind, and social perception. Functional connectivity within and between social cognitive networks was measured during a naturalistic social task. Among SSD, a significant inverse relationship was found between social cognition and cumulative metabolic burden (β=-0.38, *p*<0.001) and HA1c (β=-0.37, *p*<0.001). The relationship between social cognition and HA1c was robust across domains and measures of social cognition and after accounting for age, sex, race, non-social neurocognition, hospitalization, and treatment with different antipsychotic medications. Negative connectivity between affect sharing and motor resonance networks was a partial mediator of this relationship across SSD and HC groups (β=-0.05, *p*=0.008). There was a group x HA1c effect indicating that SSD participants were more adversely affected by increasing HA1c. Thus, we provide the first report of a robust relationship in SSD between social cognition and abnormal glucose metabolism. If replicated and found to be causal, insulin sensitivity and blood glucose may present as promising targets for improving social cognition, functional outcomes, and quality of life in SSD.

## 1 Introduction

Schizophrenia spectrum disorders (SSD) are among the leading causes of disability worldwide and take an enormous toll on patients, families, and society.^1,2^ Social cognition is a key predictor of functional impairment in people with SSD and appears to mediate the relationship between general neurocognition and functional outcomes.^3–5^ Cognitive training interventions have demonstrated efficacy in improving social cognition and functioning among individuals with SSD, but have not been widely implemented.^6–8^ Thus far, pharmacologics for social cognition show mixed results at best.^9,10^ Metabolic risk factors are common among individuals with SSD and correlate with greater impairment in general neurocognition, but their relationship with social cognition has not been evaluated.^11^ Here, we investigate the relationship between social cognition and potentially modifiable metabolic risk factors.

Key social cognitive domains include emotion processing, theory of mind, and social perception – each of which is consistently impaired across the psychosis spectrum, from the prodrome to the chronic phase of illness.^12–15^ These impairments have been tied to altered functional connectivity in social cognitive networks^16^ as well as canonical biological correlates of psychosis, for example polygenic risk,^17^ disruptions in white matter structure,^18^ and altered activity in the amygdala.^16,19^

Metabolic disturbances also appear to be integral features of SSD, with shared genetic and environmental risk factors.^20,21^ Disturbances include obesity or abdominal obesity, insulin resistance, elevated cholesterol and triglycerides, high blood pressure, and high blood glucose; the presence of multiple derangements is associated with increased risk for heart disease,^22^ the leading cause of death in SSD.^23^ Glycosylated hemoglobin or hemoglobin A1c (HA1c) is a stable measure of glucose homeostasis and insulin resistance over 120 days, with levels ≥6.5% defined as diabetes mellitus (DM), and levels ≥5.7% defined as prediabetes, indicating insulin insensitivity and increased risk for DM.^24^ While metabolic disturbances are common side effects of antipsychotic medications, blood glucose is elevated even in individuals at clinical high risk for psychosis, unaffected siblings, and unmedicated individuals in their first episode of psychosis.^25–27^

One recent meta-analysis indicated that each individual component of metabolic syndrome (i.e., hypertension, dyslipidemia, abdominal obesity/high waist circumference, and insulin resistance) was significantly associated with cognitive impairment in SSD,^28^ while another found that metabolic syndrome, hypertension and DM were significantly associated with general cognition while obesity and insulin resistance were not.^11^ Waist circumference in particular was correlated with impairment in processing speed, attention, working memory, and reasoning ^29^. The relationship between metabolic disturbance and impaired neurocognition has been widely replicated.^30–32^ While no causal link has been definitively identified, there are several plausible pathways through which metabolic disturbance may lead to decreased cognitive functioning, including alterations in vasculature, structural connectivity, and inflammation, as well as the shared effects of decreased exercise.^33–38^

Despite the finding that impairment in social cognition accounts for more variance in functional outcome than non-social neurocognition, to our knowledge, there have been no studies examining relationships between metabolic disturbance and social cognition in SSD. Here, we aimed to examine the relationship between metabolic disturbances and social cognitive impairments in SSD, compare to non-psychiatric healthy comparison subjects (HC), and investigate functional connectivity mediators and other moderators of this effect. Our main hypothesis was that we would find a significant inverse relationship between each domain of social cognition and metabolic disturbance, particularly with HA1c as a measure of insulin sensitivity and glucose intolerance.

## 2 Methods

### 2.1 Participants

The Social Processes Initiative in the Neurobiology of the Schizophrenias (SPINS) is a large multicenter study examining social cognition in SSD. Participants were assessed at the Centre for Addiction and Mental Health (CAMH), Maryland Psychiatric Research Center (MPRC), and Zucker Hillside Hospital (ZHH). Individuals with SSD met Diagnostic and Statistical Manual of Mental Disorders, 5th Edition (DSM-5)^39^ criteria for schizophrenia, schizoaffective disorder, schizophreniform disorder, or unspecified psychotic disorder. HC participants did not have a first-degree family history of psychotic disorder nor lifetime diagnosis of an Axis I psychiatric disorder with the exception of past adjustment disorder, phobic disorder, or major depressive disorder (in full remission for at least 2 years, and not on medications). Participants were interviewed using the Structured Clinical Interview for the DSM^40^ and psychiatric diagnoses were determined at an expert consensus case conference. Institutional review boards from each study site approved the study procedures. Participants provided written informed consent prior to the initiation of study procedures.

Because markedly elevated blood glucose is believed to be a confounding variable in examining brain structure, which was the primary outcome of interest in the original SPINS study, people with uncontrolled DM as reflected by a HA1c of 6.5% or greater were excluded. One HC participant was excluded from these analyses for an abnormal HA1c<4%, which may reflect a hematological disorder and would not be an accurate measure of blood sugar. Other exclusion criteria were current substance use disorder, intellectual disability, and history of traumatic brain injury.

Table 1 describes participant details. Included participants had valid results for HA1c and ≥5 of 7 social cognition tasks (*n*=28 excluded for missing values in more than two social cognition tasks).

**Table 1:** Participant Characteristics.

| Measure | SSD | HC | *p* |
| --- | --- | --- | --- |
| *n* | 245 | 165 |  |
| Site |  |  | 0.62 |
| CAMH | 84 | 49 |  |
| MPRC | 81 | 58 |  |
| ZHH | 80 | 58 |  |
| Age (years ± SD) | 32.5 ± 10.4 | 32.0 ± 10.4 | 0.71 |
| Sex (Female, *n*, %) | 81 (33%) | 75 (45%) | 0.01 |
| Race |  |  |  |
| Asian | 34 (14%) | 28 (17%) | 0.39 |
| Black | 80 (33%) | 43 (26%) | 0.15 |
| Mixed | 19 (8%) | 5 (3%) | 0.05 |
| White | 131 (53%) | 93 (56%) | 0.56 |
| Other | 5 (2%) | 0 (0%) | 0.08 |
| Ethnicity |  |  | 0.02 |
| Hispanic/Latinx | 29 (12%) | 8 (5%) |  |
| Not Hispanic/Latinx | 214 (88%) | 154 (95%) |  |
| Metabolic Measures |  |  |  |
| HA1c (mean % ± SD) | 5.4 ± 0.4 | 5.3 ± 0.4 | 0.005 |
| BMI (mean kg/m<sup>2</sup> ± SD) | 29.1 ± 5.9 | 25.9 ± 5.5 | <0.001 |
| High Waist Circumference (*n*, %) | 92 (46%) | 32 (24%) | <0.001 |
| Obese (*n*, %) | 96 (40%) | 31 (19%) | <0.001 |
| Comorbid Conditions (*n*, %) |  |  |  |
| Diabetes Mellitus, Type II | 8 (3%) | 1 (1%) | 0.07 |
| Hypertension | 17 (7%) | 1 (1%) | 0.002 |
| Hyperlipidemia | 19 (8%) | 0 (0%) | <0.001 |

*Note: P values shown for group comparisons using either t-test for continuous variables or chi-squared test for categorical variables. SSD – schizophrenia spectrum disorder; HC – healthy comparison subject; CAMH – Centre for Addiction and Mental Health; MPRC – Maryland Psychiatric Research Center; ZHH – Zucker Hillside Hospital; SD – standard deviation; HA1c – hemoglobin A1c; lbs – pounds; BMI – body mass index.*

### 2.2 Metabolic Measures

HA1c level was obtained from blood samples. Body mass index (BMI) was calculated from weight and height per the standard formula. Obesity was categorized according to the Center for Disease Control (CDC) definition of ≥30kg/m^2^.^41^ Waist circumference was measured at the superior margin of the iliac crest according to Adult Treatment Panel III (ATP III) guidelines and prior studies (high waist circumference if ≥102cm in men or ≥88cm in women).^22,42^ Measurement of waist circumference was not completed for 45 SSD and 33 HC participants. The International Diabetes Federation proposes alternate criteria for waist circumference;^43^ analysis using these criteria did not alter the results (Supplemental Materials). BMI was not able to be calculated for 6 SSD and 5 HC participants. Comorbid medical diagnoses of DM, hypertension or hyperlipidemia, as well as prescription medications to treat these disorders, were solicited from the participant’s report and verified through medical records. Because cholesterol levels were not evaluated, metabolic syndrome status could not be determined.

Cumulative metabolic burden was calculated by counting the number of the following criterion present for each participant: obesity, high waist circumference, prediabetic HA1c (≥5.7%), and diagnosis or medication for metabolic conditions (DM, hypertension, or hyperlipidemia). The main results presented were based on existing data, where only metabolic risk factors found to be present were counted and missing values were treated as being risk factors. To further substantiate our findings, we also calculated another version based on imputed data where missing values for waist circumference and BMI were estimated from available demographic and metabolic data.

### 2.3 Assessment of Cognition and Psychosis Symptoms

Neurocognition was assessed with the MATRICS consensus battery, which measures processing speed, attention and vigilance, working memory, verbal learning and memory, visual learning and memory, reasoning and problem solving, and emotional intelligence.^44^ To calculate a summary measure of non-social cognition, domain t-scores were combined into a single aggregate score with equal weighting, and a standardized z-score was calculated for the SSD sample. The MSCEIT (Mayer-Salovey-Caruso Emotional Intelligence Test) was omitted from this calculation as it is considered a measure of emotional or social intelligence. For SSD participants, overall psychosis severity was assessed with the Brief Psychiatric Rating Scale (BPRS).^45^

Social cognition was assessed with 5 tasks. The Penn Emotion Recognition 40 (ER40) captures speed and accuracy of emotion identification from photographs.^46^ An efficiency score was calculated by adding z-scores for accuracy and speed. The Empathic Accuracy (EA) task presents nine videos of actors describing emotional autobiographical events. Participants are asked to continuously rate how positive or negative the person is feeling, and participant responses were correlated with ratings from the subjects in the videos.^47^ The Reading the Mind in the Eyes Task (RMET) presents the participant with a cropped photo of the eye region and asks for identification of the corresponding mental state.^48^ Relationships Across Domains (RAD) presents a series of vignettes and asks the participant to characterize the dyadic relationship described.^49^ The Awareness of Social Inference Test-Revised (TASIT) includes 3 parts, each based on viewing brief videos showing an interaction between actors.^50^ Part 1 asks the participant to identify displayed emotions, Part 2 and Part 3 test social inference from situations involving sarcasm and lies. As each TASIT part tests a different function, they were considered separately in the analyses. Individual task scores were standardized across the whole sample after undergoing Yeo-Johnson transformation to adjust for left-skewness.^51^

Three social cognition domains were represented.^52^ The Emotion Processing domain consisted of ER40, EA, and TASIT Part 1. The Theory of Mind domain consisted of RMET and TASIT Parts 2 and 3. The Social Perception domain consisted of the RAD. Domain scores were calculated by summing task scores and re-standardizing across the whole sample. An overall social cognition score included all tasks. Even weights were used for domain and overall scores. Weighted single- and two-factor scores for social cognition based on previously published factor analyses^5^ were also explored and results were consistent with that of the overall social cognition and domain scores reported here.

There were missing or invalid scores for 7 participants for ER40, 3 participants for TASIT Part 3, 1 participant for RMET, and 5 participants for RAD. To further test the validity of our conclusions, we repeated the analyses after using multiple imputation to address these missing values.

### 2.4 Functional Connectivity of Social Cognitive Networks

Magnetic resonance imaging (MRI) scans were collected on five 3T scanners and harmonized as detailed previously.^53^ The EA task^47^ was completed during functional MRI. Detailed methods and calculation of connectivity measures during the EA task were described previously by Oliver et al.^53^ Briefly, state-related background connectivity analysis was performed for critical regions of interest associated with social cognition and social cognitive networks were identified using the Louvain community detection algorithm for signed networks. Within- and between-positive and negative connectivity strengths were derived and standardized for the three identified networks: *motor resonance* (inferior frontal gyrus, premotor cortex, supplemental motor area, inferior parietal lobule, posterior superior temporal sulcus), *affect sharing* (anterior insula, anterior cingulate cortex, and regions of the inferior parietal lobule, and *mentalizing* (medial prefrontal cortex, anterior cingulate, temporal parietal junction, posterior superior temporal sulcus, and precuneus). These network measures are available for a subset of the participants (*n*=164 SSD, *n*=117 HC) collected prior to a scanner upgrade at the CAMH site.

### 2.5 Statistical Analyses

All analyses were conducted using R version 3.5.2.^54^ Potential outliers in overall social cognition performance and HA1c level were separately evaluated in the SSD and HC groups, using the adjusted boxplot and standard interquartile range methods.^55^ Influence of the potential outliers was assessed with Mahalanobis distance using the default threshold of α=0.025 and the *performance* package in R.^56^ There were no influential outliers.

Multiple linear regressions were used to evaluate the relationships between social cognition and metabolic predictors as well as potential demographic and clinical covariates (Supplemental Table 1 & 2). Standardized β coefficients are reported. Fisher Z-transformation was used to identify 95% confidence intervals (95%CI) for key correlation coefficients. For dichotomous predictors (obesity, waist circumference), effect size was calculated with Cohen’s *d*, and significance was tested with ANOVA. Alpha was two-sided and set at 0.05. Multiple imputation was performed on missing metabolic and social cognition data using the *mice* package in R^57^ and the predictive mean matching method, which is robust to transformations.

Mediation analyses were performed using the lavaan package in R,^58^ including multiple regressions with HA1c as a predictor of overall social cognition and each of the six functional connectivity measures as mediators. Mediation models were tested using full information maximum likelihood estimation. Indirect effects were determined using nonparametric bootstrapping with 5000 resamples, where the effect is considered significant (*p*<0.05) if the bias-corrected and accelerated 95% confidence interval (CI) does not include 0.^59^

## 3 Results

### 3.1 Metabolic Burden and Social Cognition in SSD

There was a significant relationship between increased metabolic burden and greater impairment in overall social cognition (Figure 1; β=-0.38, *p*<0.001). As detailed in Supplemental Table 1, this effect remained significant after covarying for age, sex, race, and ethnicity (β=-0.26, *p*=0.002), and after covarying for overall psychosis severity as measured by the BPRS total score (β=-0.38, *p*<0.001), but became trend-level when covarying for non-social neurocognition (β=-0.11, *p*=0.08). When using imputed values for missing data, the relationship between cumulative metabolic burden and overall social cognition was significant on its own (β=-0.33, *p*<0.001), but became trend-level after covariation with demographic variables (β=-0.13, *p*=0.06) or age and non-social neurocognition (β=-0.09, *p*=0.10).

**Figure 1.**
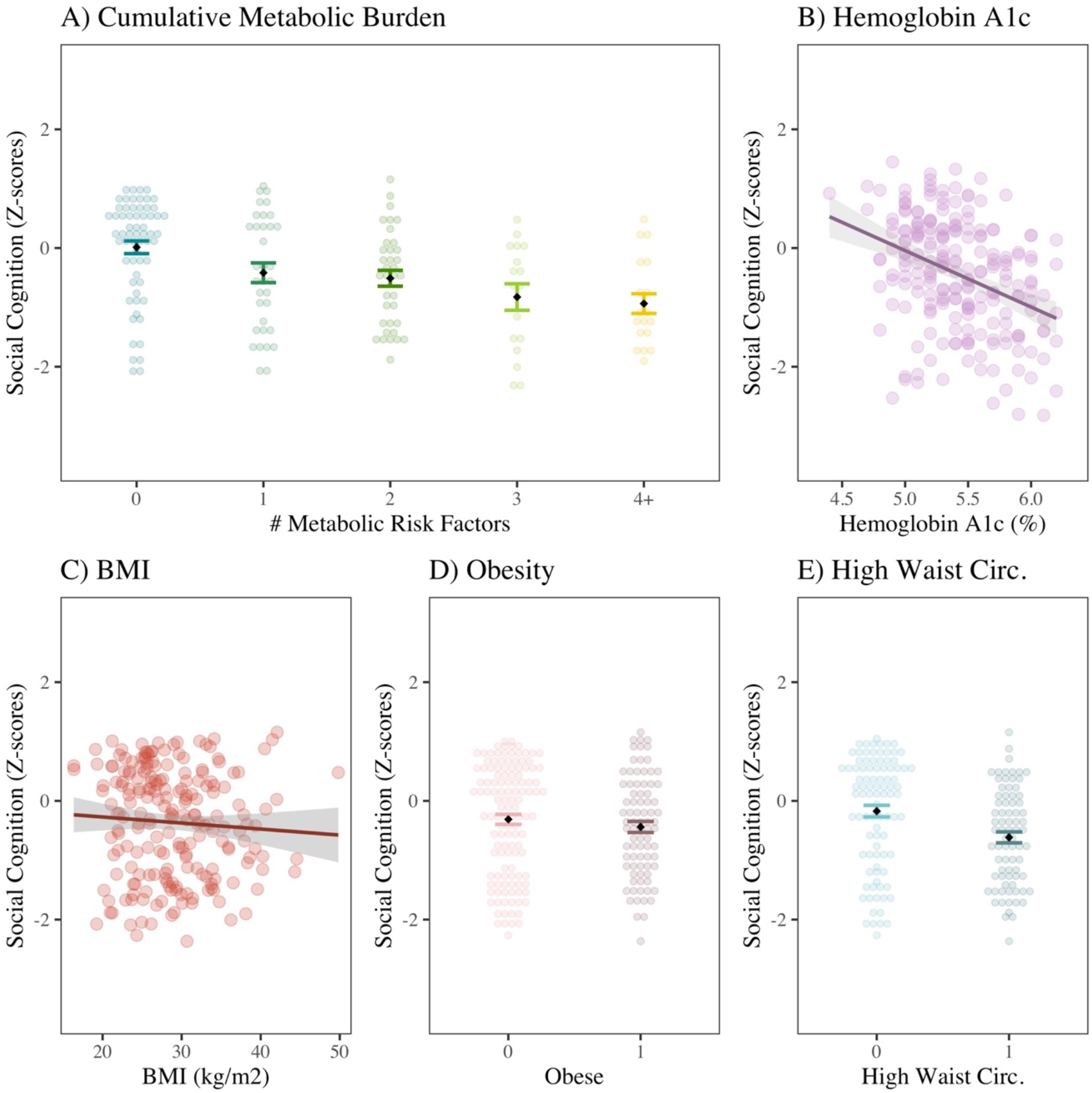
Social Cognition and Metabolic Risk in SSD: (A) Cumulative metabolic burden included the following: obesity, high waist circumference, prediabetic HA1c, diagnosis or medication for DM, diagnosis or medication for hypertension, and diagnosis or medication for hyperlipidemia; on a linear regression, there was a significant relationship between higher number of metabolic risk factors and greater impairment in social cognition (β=-0.38, *p*<0.001, Adj R^2^=0.14). B) There was a significant negative relationship between overall social cognition and HA1c (β=-0.37, *p*<0.001, Adj R^2^=0.14). There was no significant relationship between social cognition and (C) Body Mass Index (β=-0.05, *p*=0.49) or (D) Obesity (Cohen’s *d*=0.09, *p*=0.34). (E) High waist circumference was significantly correlated with greater impairment in social cognition (Cohen’s *d*=0.49, *p*=0.002). However, only HA1c remains significantly correlated with social cognition after covarying for age, sex, race, ethnicity, and non-social neurocognition (β=-0.16, *p*=0.005, Adj R^2^=0.53). Standardized β coefficients are reported.

The individual metabolic risk factors were also evaluated. Hemoglobin A1c (HA1c) was significantly correlated with overall social cognition (β=-0.37, 95% CI [-0.47, -0.26], *p*<0.001); this finding was robust despite covarying for non-social neurocognition and demographic variables (β=-0.16, *p*=0.005), overall psychosis severity (β=-0.38, *p*<0.001), and duration of illness (β=-0.27, *p*<0.001). The association between HA1c and social cognition remained significant when using imputed values for missing social cognition task results both as a simple linear regression (β=-0.35, *p*<0.001) and after covarying for demographic variables and non-social neurocognition (β=-0.13, *p*=0.01). High waist circumference was also significantly associated with greater impairment in social cognition (Cohen’s *d*=0.49, *p*=0.002). Neither BMI nor obesity were significantly correlated with overall social cognition.

As a single measure, HA1c accounted for a similar amount of variance in overall social cognition (R^2^=0.14) as cumulative metabolic burden (R^2^=0.14). HA1c remained significantly correlated with social cognition even after covarying for non-social neurocognition, and results for were consistent when using imputed values. The effect size for the correlation between HA1c and social cognition (β=-0.38) was greater than that of overall psychosis severity (β=-0.15). Because of the strength of this relationship, HA1c was the focus of additional characterization and exploration.

### 3.2 Hemoglobin A1c and Social Cognition in SSD

HA1c was negatively related to each of the three social cognition domains: Emotion Processing (Figure 2; β=-0.36, *p*<0.001), Theory of Mind (β=-0.31, *p*<0.001), and Social Perception (β=-0.27, *p*<0.001). The correlations remained significant for Emotion Processing and Theory of Mind after covarying for demographic variables and non-social neurocognition, but became trend-level for Social Perception after covarying for demographic variables (Supplemental Table 2). Using imputed values for missing social cognition data did not alter the relationship between HA1c and the social cognition domains.

**Figure 2.**
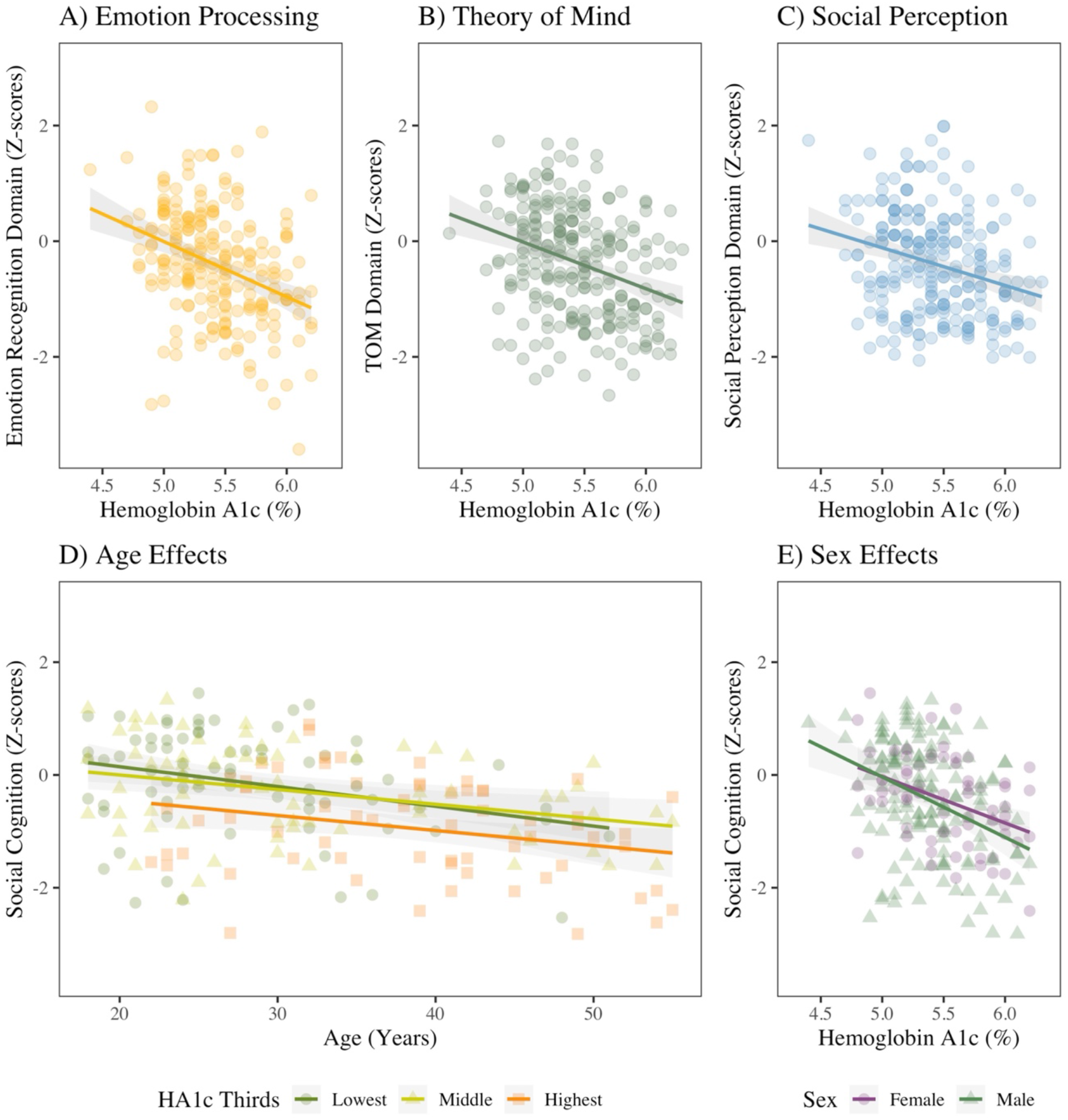
Hemoglobin A1c and Social Cognition in SSD: Among participants with schizophrenia spectrum disorders (SSD), there was a significant inverse relationship between hemoglobin A1c and each domain of social cognition: (A) Emotion Processing (β=-0.36, *p*<0.001), (B) Theory of Mind (β=-0.31, *p*<0.001), and (C) Social Perception (β=-0.27, p<0.001). (D) Age was also related to greater impairment in aggregate social cognition (β=-0.40, *p*<0.001), but elevated HA1c in the top third of the SSD group (≥5.7%) was related to still greater impairment (β=-0.25, *p*=0.001). (E) There was no significant effect of sex on the relationship between HA1c and aggregate social cognition (β=0.01, *p*=0.93). Standardized β coefficients are reported.

There was a significant interaction with age, which was also correlated with greater impairment in social cognition (Figure 2D; Supplemental Table 1 & 2). There was no interaction with sex (Figure 2E; Supplemental Table 1 & 2).

All individual social cognition tasks were inversely correlated with HA1c (Supplemental Figure 1). The strongest relationships were with the TASIT Part 1 and EA tasks, each with β=-0.31, *p*<0.001, R^2^=0.09. The weakest relationship was with the ER40, β=-0.19, *p*=0.003, R^2^=0.03. Each part of the TASIT, the RMET, and the EA scores remained significantly inversely correlated with HA1c even after covarying for key demographic variables and non-social neurocognition, but ER40 and RAD were no longer significantly associated.

### 3.3 Group by Hemoglobin A1c Effect on Social Cognition

We next examined whether there was a significant group effect on the relationship between social cognition and HA1c and whether the relationship existed for HC participants (Figure 3). Overall, the HC group performed better on the social cognition tasks than the SSD group. There was a significant group by HA1c interaction for overall social cognition, Emotion Processing, and Social Perception domains, where higher levels of HA1c were associated with greater impairment for the SSD group. In an examination of the HC group alone, there was also a significant inverse relationship between social cognition and HA1c (β=-0.20, 95%CI [-0.34, -0.05], *p*=0.01), but the correlation was no longer significant after covarying for demographic variables or non-social neurocognition, perhaps due to less power and lower effect size in HC.

**Figure 3.**
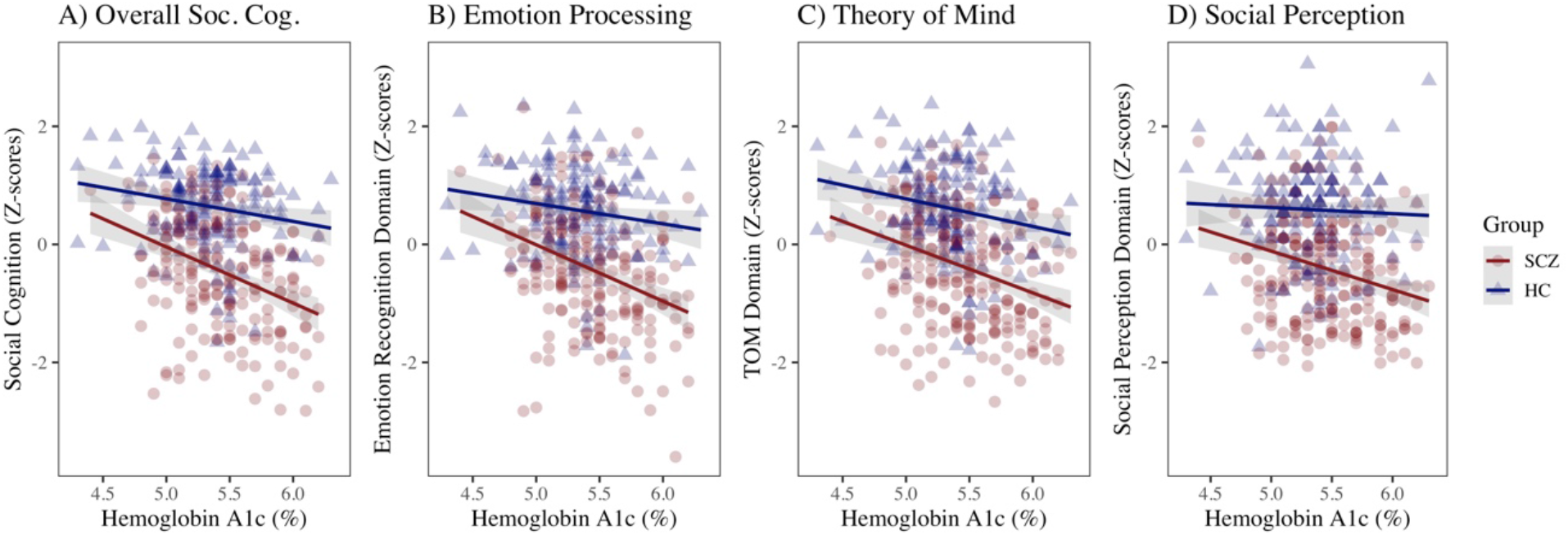
SSD Group by Hemoglobin A1c Effect on Social Cognition: (A) Overall Social Cognition, HA1c (β=-0.35, *p*<0.001), HC group (β=-1.0, *p*=0.10), and HC group x HA1c (β=1.5, *p*=0.01). (B) Emotion Processing Domain, HA1c (β=-0.35, *p*<0.001), HC group (β=-1.2, *p*=0.07), and HC group x HA1c (β=1.6, *p*=0.01). (C) Theory of Mind Domain, HA1c (β=-0.30, p<0.001), HC group (β=-0.45, *p*=0.47), and HC group x HA1c (β=0.90, *p*=0.15). (D) Social Perception Domain, HA1c (β=-0.24, *p*<0.001), HC group (β=-0.99, *p*=0.12), and HC group x HA1c (β=1.45, *p*=0.02). Standardized β coefficients are reported.

### 3.4 Functional Connectivity Mediators for Hemoglobin A1c and Social Cognition

Functional connectivity measures during the EA task were explored as potential mediators of the relationship between HA1c and overall social cognition. Within- and between-network positive and negative connectivity for motor resonance, affect sharing, and mentalizing networks were included in the analyses. Of these, negative connectivity between the affect sharing and motor resonance networks was found to be a significant partial mediator (β (indirect)= -0.05, 95%CI [-0.22, -0.04], p=0.008; Figure 4).

**Figure 4.**
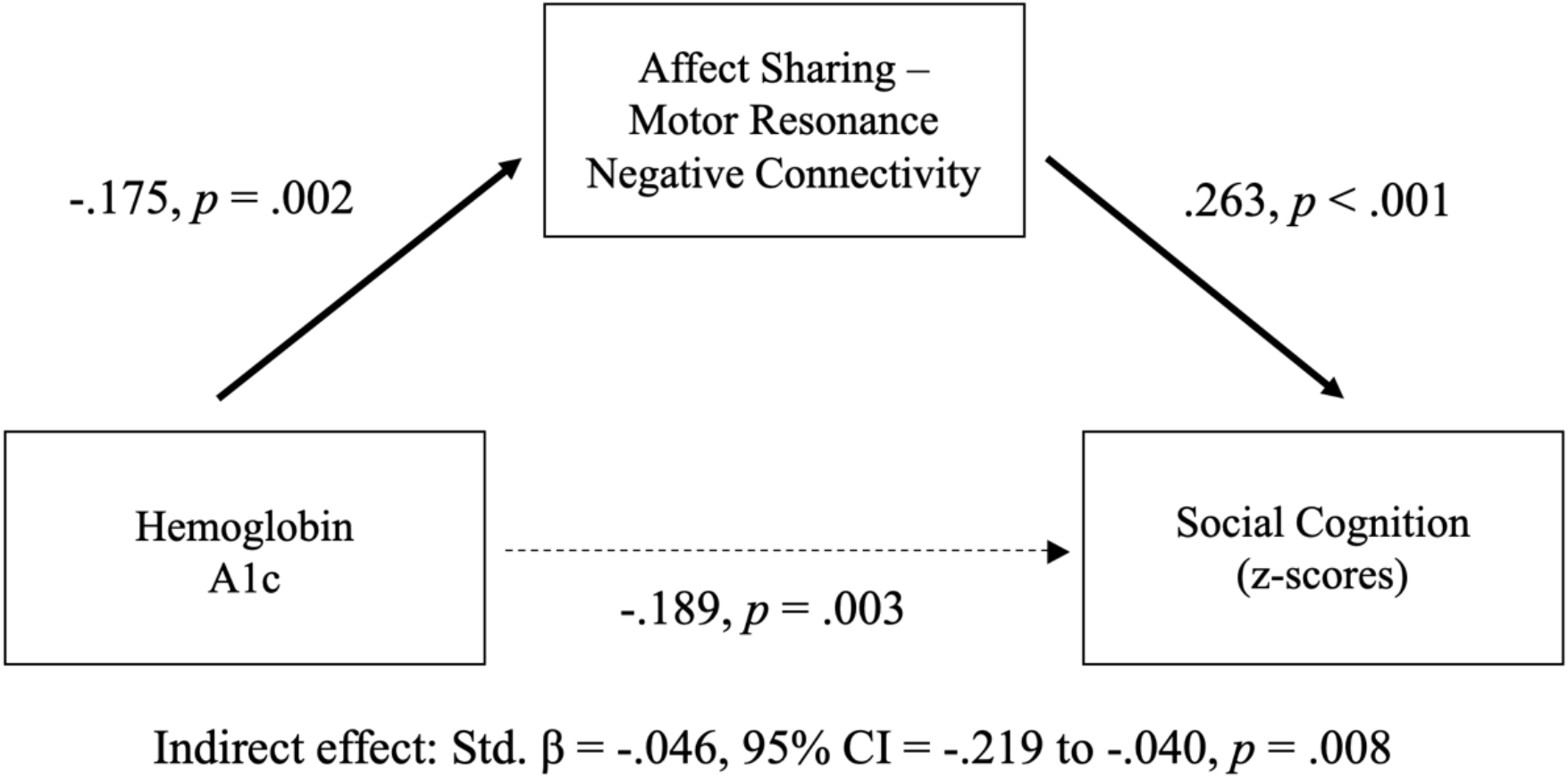
Functional Connectivity Mediation of Hemoglobin A1c and Social Cognition Across SSD and HC Groups: The association between hemoglobin A1c and overall social cognition was partially mediated by negative connectivity between the affect sharing and motor resonance networks (*n*=253).

### 3.5 Hospitalization and Antipsychotic Medications, and Effect on Social Cognition

To assess whether treatment history impacted the relationship, we explored the effect of number of hospitalizations and treatment with antipsychotic medications on the relationship between HA1c and social cognition (Supplemental Figure 2). There was no relationship between the number of hospitalizations reported by the participant and overall social cognition. Treatment with first-or second-generation antipsychotics did not have a significant effect on the inverse relationship between HA1c and social cognition.

The effect of individual antipsychotic medications on the relationship between HA1c and social cognition was also explored for participants reporting use of aripiprazole (*n*=26), clozapine (*n*=57), risperidone or paliperidone (*n*=54), and olanzapine (*n*=25). Other medications were not used with sufficient frequency to allow for analysis. The inverse relationship between HA1c and social cognition remained significant despite use of any of these medications. Treatment with aripiprazole was associated with preservation of social cognition (β=0.13, *p*=0.047).

### 3.6 Non-Social Neurocognition and Hemoglobin A1c in SSD

The relationship between non-social neurocognition and HA1c was also explored (Supplemental Figure 3). Aggregate non-social cognition (β=-0.11, 95%CI [-0.23, 0.02], *p*=0.10), and the domains of processing speed, working memory, attention and vigilance, and reasoning were not significantly correlated with HA1c. However, both verbal (β=-0.22, *p*<0.001) and visual learning (β=-0.22, *p*<0.001) were significantly negatively associated with HA1c. Neither of these associations remained significant after covarying for demographic variables (age, sex, race, and ethnicity).

## 4 Discussion

To our knowledge, this is the first study examining metabolic burden or blood glucose and social cognition in SSD. We found a significant relationship between metabolic burden and impairment in social cognition. HA1c predicted a similar amount of variance in overall social cognition as cumulative metabolic burden and was more robust when accounting for covariates. BMI and obesity were not related to social cognition, but abdominal obesity was associated with greater impairment. The relative importance of HA1c for cognition compared to other metabolic disturbances is consistent with prior studies in the non-psychiatric population where the relationship between hyperglycemia and cognitive impairment is well-replicated while the relationship between metabolic syndrome and cognitive impairment is more heterogeneous.^60^ Therefore, HA1c was prioritized for further exploration in our subsequent analyses.

We found that increased Ha1c was significantly correlated with impaired functioning in each social cognition domain, with more robust effects on emotion processing and theory of mind than social perception. Decreased negative connectivity between the affect sharing and motor resonance functional networks partially mediated the relationship between HA1c and overall social cognition. Of the measures tested, it is not surprising that negative connectivity between the affect sharing and motor resonance networks should emerge as the one significant mediator as this was the only functional measure that was significantly correlated with both emotion processing and theory of mind factors in a previous study^53^ and thus may represent functional changes that are particularly important for social cognitive performance. The relationship between blood glucose and functional connectivity has not been previously reported for SSD, to our knowledge, but alterations in resting-state functional connectivity have been found for individuals with DM.^61,62^

The current study does not provide information on causality in these relationships. However, there are several mechanistic pathways that could plausibly explain the relationship between HA1c and impaired functional connectivity, and subsequent decrease in social cognitive performance. Chronically elevated blood glucose and metabolic disturbance result in large vessel and microvascular changes, which profoundly alter cerebral circulation and brain structure.^33,63^ For example, previous studies have found an association between higher fasting glucose or prediabetes and decreased fractional anisotropy in white matter tracts,^34,35^ as well as between metabolic syndrome and hypoperfusion of the left orbital prefrontal cortex.^36^ Inflammation may play a role through elevation of interleukin-6 and C-reactive protein.^64,65^ Elevated blood sugar can also lead to neuronal dysfunction through oxidative stress, accumulation of aberrant metabolites, and depletion of vital cofactors.^37^ Alternatively, it is possible that poor social cognition may affect health-related behaviors such as diet and exercise, which lead to metabolic disturbance, higher HA1c, and brain changes. Or, it is also possible that SSD is associated with increased inflammation, which causes both metabolic disturbance and impairments in social cognition.^31^

Our results also suggest that for SSD, the correlation of HA1c with social cognition is stronger than for non-social cognition. The effect sizes we found for HA1c and non-social cognition are aligned with findings in the non-psychiatric population where Type 2 DM explains about 10% of variance in cognition.^66^ In SSD, some studies on non-social neurocognition and metabolic risk factors have been inconsistent despite the overall evidence suggesting an inverse relationship.^28,67,68^ Consistent with our results, verbal and visual learning were related to glucose disturbance or diabetes in several prior studies.^28,34,69^ Furthermore, there was a significant group by HA1c effect such that higher HA1c was related to greater impairments in overall social cognition for SSD than for HC. The posited disruption in brain connectivity secondary to elevated blood glucose may explain this finding. Because structural and functional connectivity are already disrupted in SSD, this may leave individuals more vulnerable to further disturbances caused by elevated blood glucose, thereby resulting in the group x HA1c effect.

Examining potential moderators, the inverse correlation between social cognition and HA1c was not affected by sex, number of hospitalizations, or treatment with first-or second-generation antipsychotic medication. Both age and duration of illness were correlated with greater impairment in social cognition, whereas treatment with aripiprazole was associated with decreased impairment. Our finding that increased age is an additive risk for social cognitive impairments fits with existing literature. Though there are no reports of age-related decline in social cognition per se in SSD, general neurocognition has been observed to decline with age in SSD.^70,71^ In addition, social cognition has been observed to decline with age in the typically aging population.^72–75^ To our knowledge, no prior study has reported that aripiprazole was associated with less impairment in social cognition. This relationship may reflect the properties of the drug itself (in particular, partial agonism at dopamine receptors) or that aripiprazole may be used more frequently in people who are younger, earlier in the course of their illness or have less severe forms of the illness.

The SPINS dataset was designed as a dimensional approach to characterize social processes, including neurocircuitry correlates. For that reason, individuals with uncontrolled DM were excluded from the study due to concerns about potential confounding of the imaging findings. The study parameters also preclude any conclusions about causality. These are notable limitations to the current study. Other components of metabolic syndrome, including lipids and blood pressure, were not reliably measured. Therefore, it was not possible to determine whether participants met the criteria for metabolic syndrome. We relied on patient report and chart review as indicators of metabolic conditions, which likely underestimates the true prevalence of metabolic conditions in SSD, as these conditions tend to be underdiagnosed and undertreated in this population.^42^ As such, the cumulative metabolic burden should not be interpreted as reflecting the absolute metabolic burden experienced by participants – rather, the significance lies with the correlation between social cognition impairment and *relatively* increased metabolic burden. Additional measures of insulin sensitivity may also be informative in future studies. We attempted to account for the effect of antipsychotic medications on the reported results. However, medication dosages were not always available and there is no established method for comparing metabolic potency between different antipsychotic medications. When examining the contributions of individual medications, sample sizes became relatively small, and therefore results should be considered exploratory. Structural connectivity of white matter tracts in social cognitive networks would also be an important potential mediator to explore in future studies.

Nevertheless, our findings may have significant clinical implications. Impairment in social cognition is a key contributor to functional impairment and poor quality of life in schizophrenia.^3,4^ If insulin sensitivity or blood glucose were to play a causal role in social cognitive impairment, these could be novel modifiable targets for improving outcomes in schizophrenia. Pharmacological and non-pharmacological interventions for lowering blood glucose could be applied to improve social cognition and functioning. It is particularly notable that the range of HA1c examined in this study is limited to the non-diabetic (or well-controlled) range. Individuals with HA1c in this range would normally not come to medical attention for their blood glucose levels. It may be the case that the risk/benefit assessment in SSD would lead to acrecommendation for a lower target HA1c than in the non-psychiatric population, much in the same way that a lower cholesterol level is targeted in individuals with other risk factors for heart disease. Further studies, and particularly longitudinal studies, are critical to determining whether metabolic disturbance or HA1c play a causal role in social cognition.

***4*.*1 Conclusion***

In this study, we provide the first report of a robust and significant relationship in SSD between social cognition and metabolic burden, particularly HA1c – as a measure of glucose metabolism and insulin sensitivity. The relationship was partially mediated by decreased functional connectivity between social cognition networks and was consistent across different measures and domains of social cognition and after accounting for age, sex, race, non-social neurocognition, hospitalization, and treatment with different antipsychotic medications. If replicated and found to be causal, blood glucose may present as a promising target for improving social cognition, functional outcomes, and quality of life in SSD.

## Data Availability

All data produced in the present study are available upon reasonable request to the authors.

## Acknowledgements

We thank the participants for their contribution to this work. In addition, we thank Kristin Minara from Zucker Hillside Hospital. Funding was received from NIMH, R01MH102313.

## SUPPLEMENTAL MATERIALS

**Supplemental Table 1:**
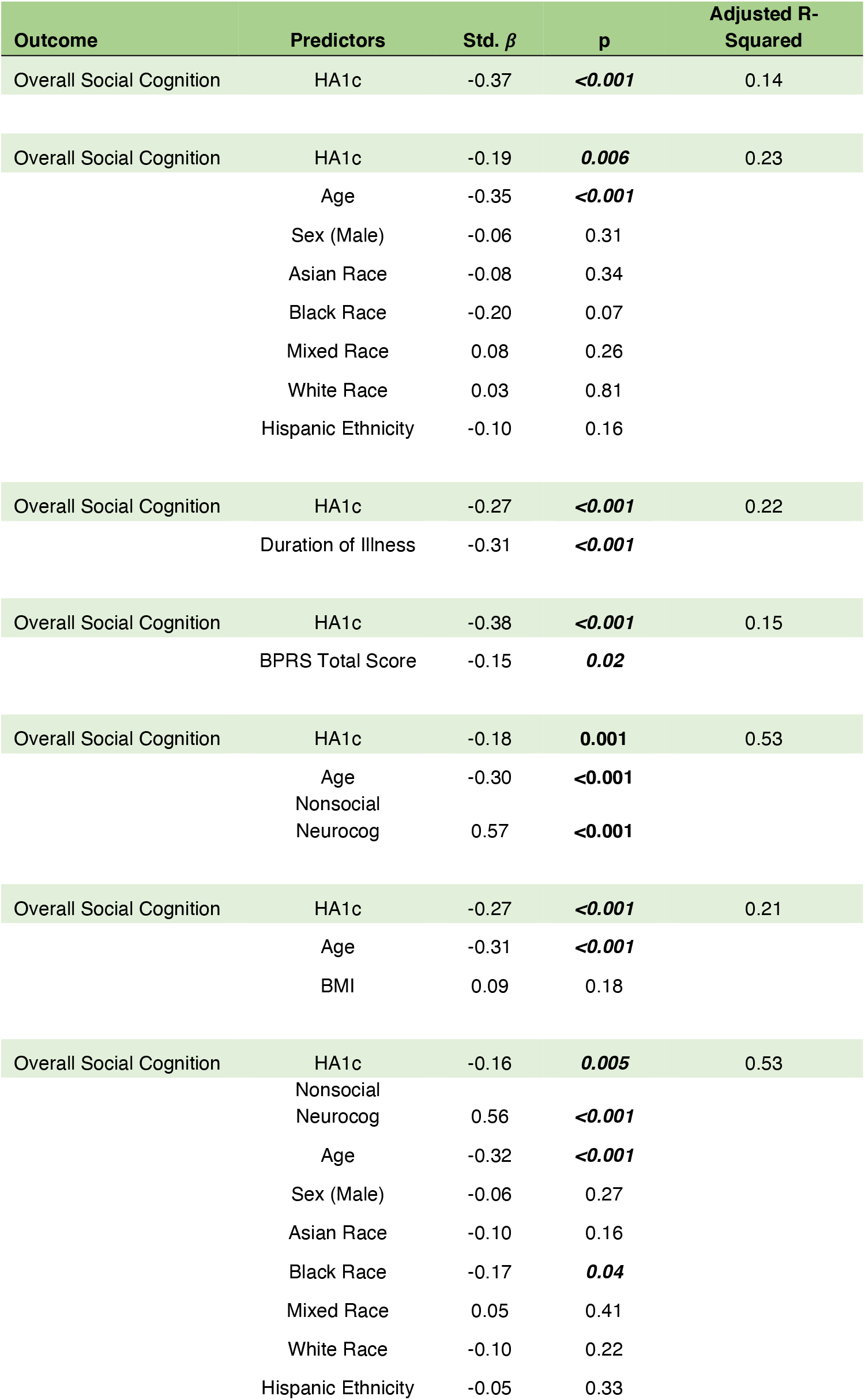
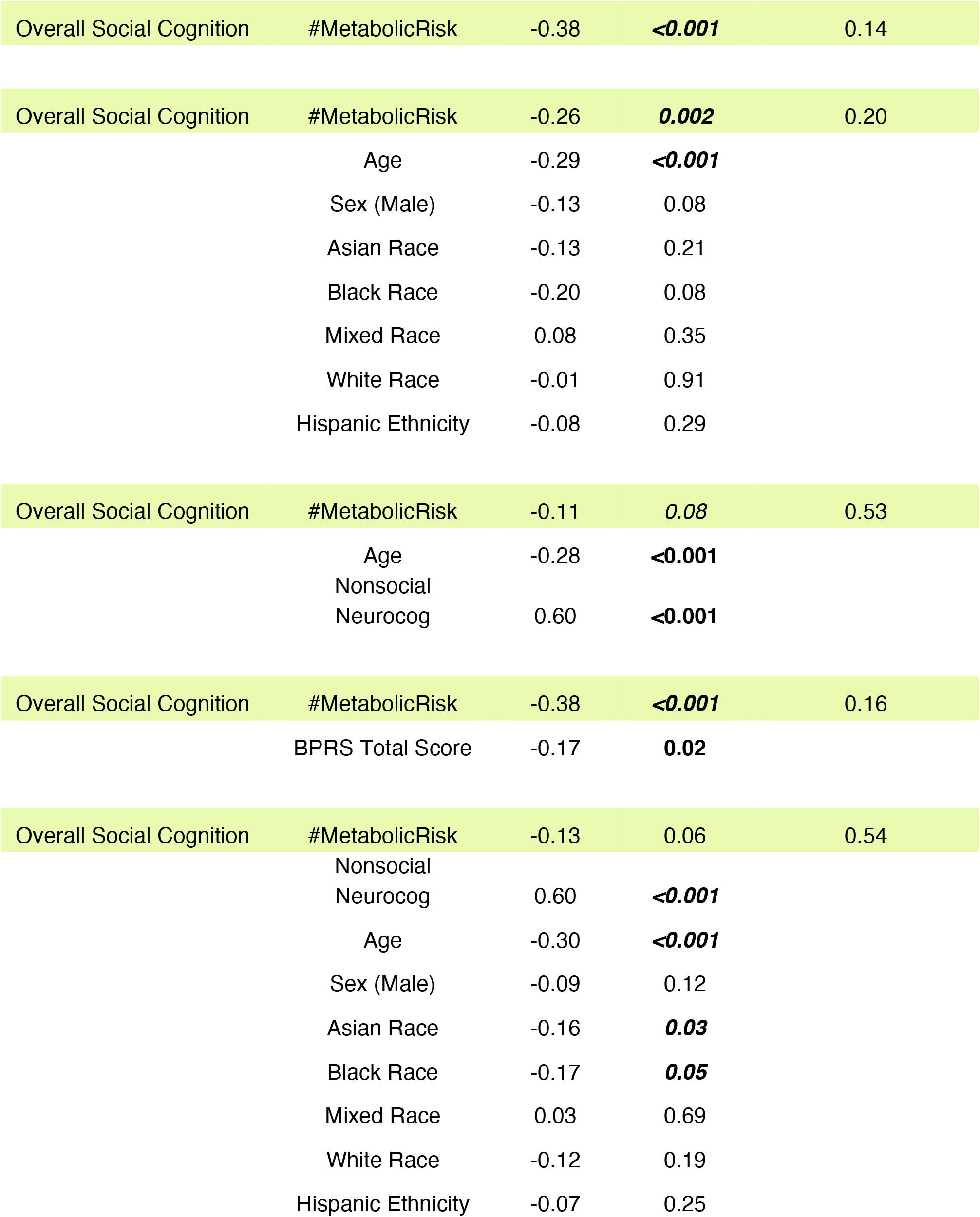
Overall Social Cognition, Metabolic Risk, and Hemoglobin A1c.

**Supplemental Table 2:** Social Cognition Domains and Hemoglobin A1c.

| Outcome | Predictors | Std. $\beta$ | $p$ | Adjusted R-Squared |
| --- | --- | --- | --- | --- |
| Emotion Processing Domain | HA1c | -0.36 | **<0.001** | 0.13 |
| Emotion Processing Domain | HA1c | -0.16 | **0.01** | 0.27 |
|  | Age | -0.43 | **<0.001** |  |
|  | Sex (Male) | -0.08 | 0.18 |  |
|  | Asian Race | 0.02 | 0.84 |  |
|  | Black Race | -0.05 | 0.65 |  |
|  | Mixed Race | 0.03 | 0.69 |  |
|  | White Race | 0.14 | 0.2 |  |
|  | Hispanic Ethnicity | -0.03 | 0.65 |  |
| Emotion Processing Domain | HA1c | -0.14 | **0.01** | 0.47 |
|  | Age | -0.39 | **<0.001** |  |
|  | Nonsocial Neurocog | 0.47 | **<0.001** |  |
| Theory of Mind Domain | HA1c | -0.31 | **<0.001** | 0.09 |
| Theory of Mind Domain | HA1c | -0.22 | **0.002** | 0.11 |
|  | Age | -0.16 | **0.02** |  |
|  | Sex (Male) | -0.06 | 0.37 |  |
|  | Asian Race | -0.14 | 0.14 |  |
|  | Black Race | -0.23 | **0.04** |  |
|  | Mixed Race | 0.12 | 0.11 |  |
|  | White Race | -0.09 | 0.42 |  |
|  | Hispanic Ethnicity | -0.15 | **0.03** |  |
| Theory of Mind Domain | HA1c | -0.18 | **0.001** | 0.40 |
|  | Age | -0.14 | **0.01** |  |
|  | Nonsocial Neurocog | 0.55 | **<0.001** |  |
| Social Perception Domain | HA1c | -0.27 | **<0.001** | 0.07 |
| Social Perception Domain | HA1c | -0.12 | 0.07 | 0.16 |
|  | Age | -0.22 | **0.001** |  |
|  | Sex (Male) | -0.02 | 0.75 |  |
|  | Asian Race | -0.23 | **0.01** |  |
|  | Black Race | -0.40 | **<0.001** |  |
|  | Mixed Race | 0.18 | **0.01** |  |
|  | White Race | -0.15 | 0.2 |  |
|  | Hispanic Ethnicity | -0.22 | **0.001** |  |
| Social Perception Domain | HA1c | -0.14 | **0.02** | 0.33 |
|  | Age | -0.17 | **0.004** |  |
|  | Nonsocial Neurocog | 0.50 | **<0.001** |  |

**Supplemental Figure 1.**
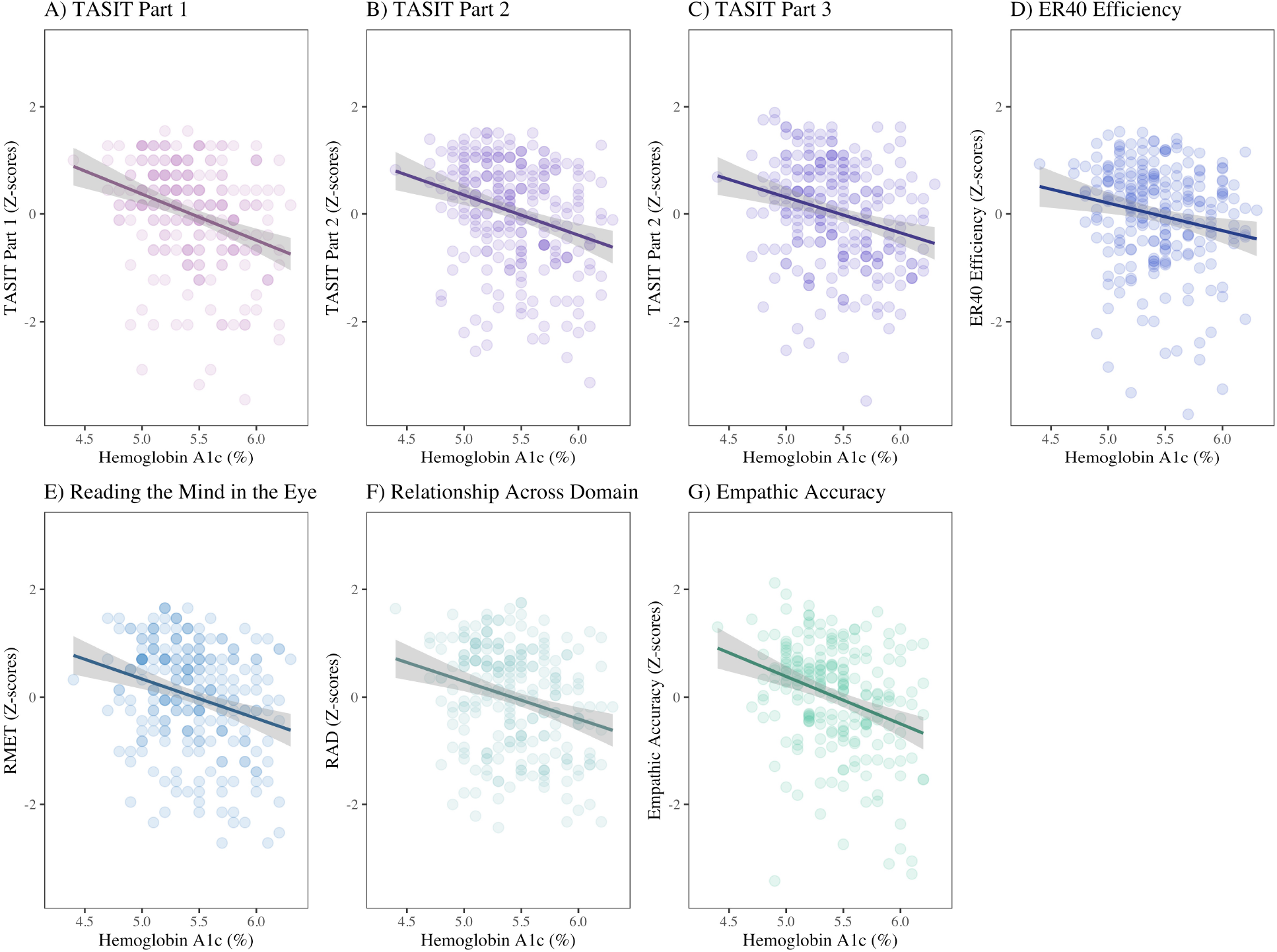
Individual Social Cognition Tasks and Hemoglobin A1c: Relationships between standardized z-scores for each social cognition task and HA1c level are shown. (A) The Awareness of Social Inference Task (TASIT) Part 1, β=-0.31, *p*<0.001, R^2^=0.09. (B) TASIT Part 2, β=-0.28, *p*<0.001, R^2^=0.08. (C) TASIT Part 3, β=-0.26, *p*<0.001, R^2^=0.07. (D) Penn Emotion Recognition 40 (ER40) Efficiency, β=-0.19, *p*=0.003, R^2^=0.03. (E) Reading the Mind in the Eyes Test (RMET), β=-0.27, *p*<0.001, R^2^=0.07. (F) Relationship Across Domains Task (RAD), β=-0.27, *p*<0.001, R^2^=0.07. (G) Empathic Accuracy (EA), β=-0.31, *p*<0.001, R^2^=0.09. Standardized β coefficients are reported.

**Supplemental Figure 2.**
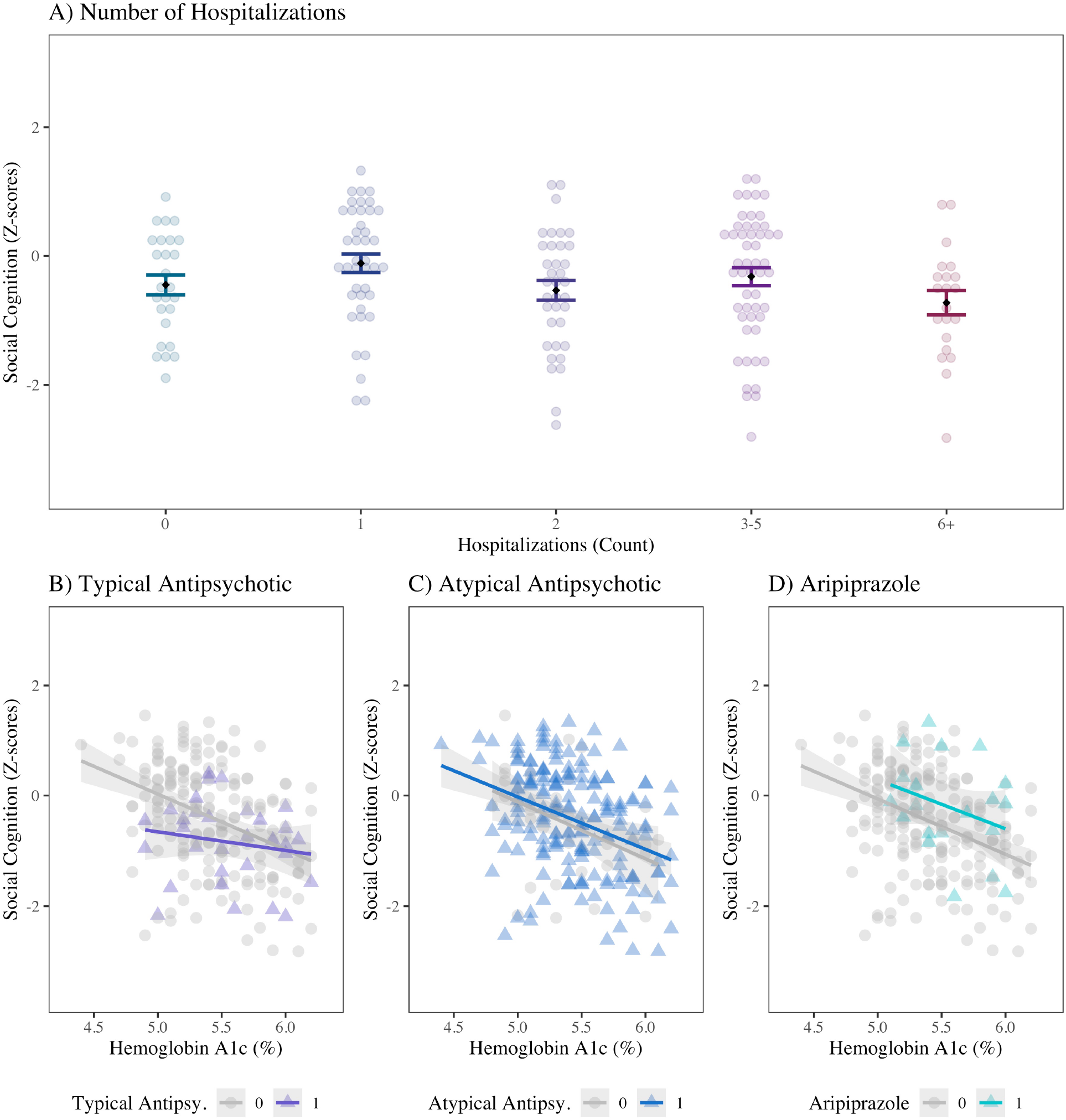
Hospitalization and Antipsychotic Treatment Effects: (A) Number of prior hospitalizations was not related to aggregate social cognition scores (β=0.10, *p*=0.16). (B) Treatment with typical antipsychotic medications had a trend level effect on the relationship between HA1c and social cognition (β=0.12, *p*=0.055), but the negative relationship between HA1c and social cognition remained highly significant. (C) Treatment with an atypical antipsychotic (β=0.11, *p*=0.47) did not significantly affect the relationship between HA1c and overall social cognition score. (D) Treatment with aripiprazole was associated with significant increase in overall social cognition score (β=0.13, *p*=0.047) but did not disrupt the correlation between HA1c and social cognition. Not shown, we also examined clozapine and olanzapine together as one group, given the similarity of their metabolic effects. There was no significant effect of treatment with either clozapine or olanzapine on the relationship between HA1c and social cognition (β=-0.09, *p*=0.17). Standardized β coefficients are reported.

**Supplemental Figure 3.**
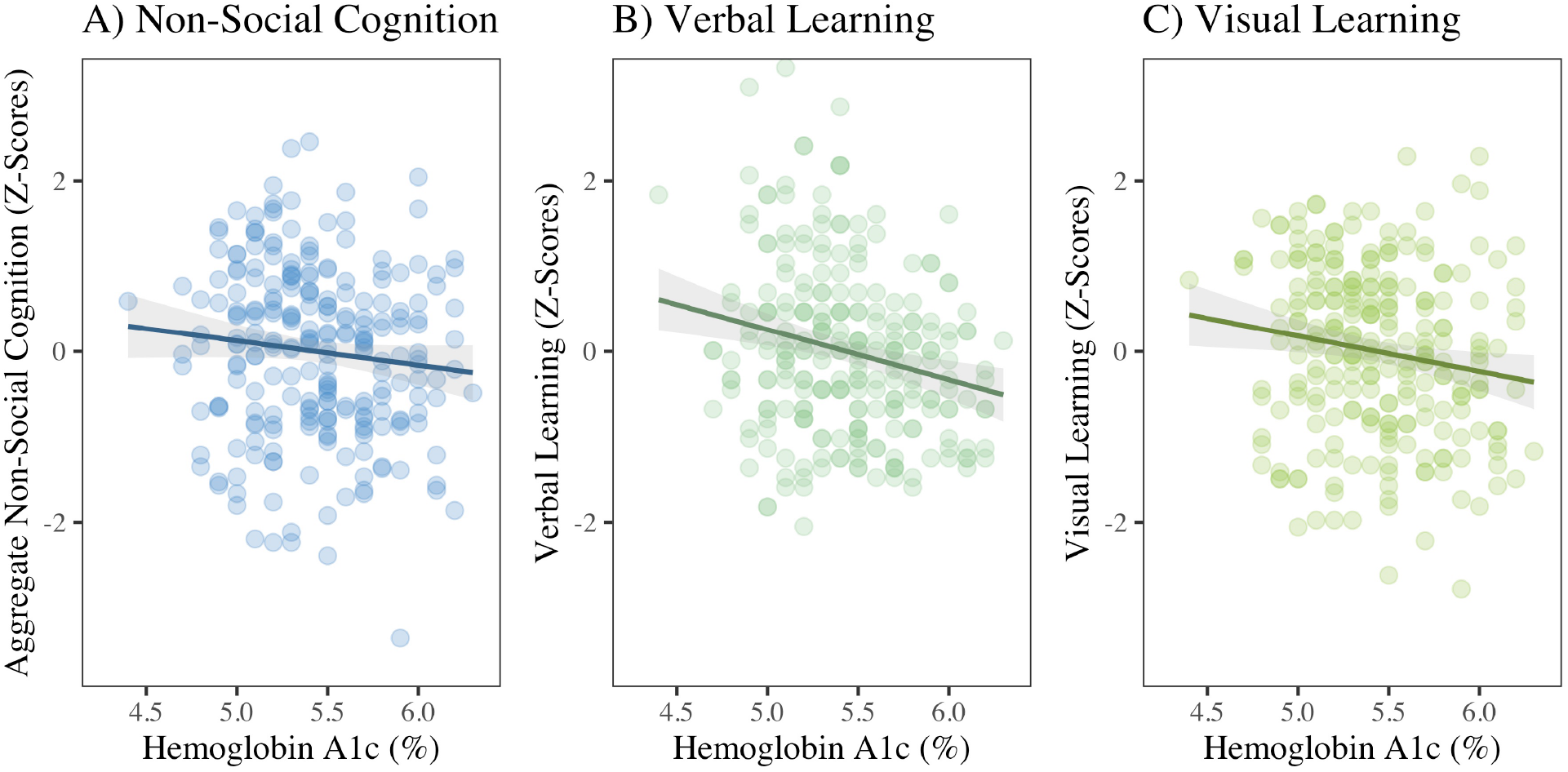
Non-social Neurocognition and Hemoglobin A1c in SSD: (A) Aggregate non-social neurocognition was not correlated with HA1c (β=-0.11, *p*=0.10). (B) Verbal learning and memory was significantly related to HA1c (β=-0.22, *p*<0.001). (C) Visual learning and memory was significantly related to HA1c (β=-0.16, *p*=0.01). There was no significant relationship between HA1c and processing speed, attention and vigilance, working memory, or reasoning. Standardized β coefficients are reported.

## Supplemental Information on Waist Circumference

The International Diabetes Federation (IDF) defines separate waist circumference cutoff points for individuals of Asian descent (Chinese, Japanese, and South Asian): 90cm for men and 80cm for women vs. individuals of European and other descent: 94cm for men and 80cm for women.^43^

Analysis using these criteria did not alter our main findings.

Overall social cognition continued to be more impaired among individuals with high waist circumference (Cohen’s *d*=-0.36, *p*=0.04).

Cumulative metabolic risk, re-calculated using the IDF guidelines, continued to be significantly correlated with greater impairment in overall social cognition (β=-0.37, *p*<0.001). As before, the results remain significant when covarying for sex, race, and ethnicity (β=-0.24, *p*=0.005), but are no longer significant when covarying for nonsocial neurocognition.

